# Multiple Physiological and Behavioural Parameters Identification for Dietary Monitoring Using Wearable Sensors: A Study Protocol

**DOI:** 10.1101/2024.06.19.24309180

**Authors:** Mayue Shi, Jiaying Zhou, Mingzhu Cai

**Author notes:** Correspondence to Dr Mingzhu Cai, and Dr Mayue Shi,. These authors contributed equally to this work (Mayue Shi, Mingzhu Cai).

## Abstract

**Introduction:** Traditional dietary intake assessment is labour- and time-consuming and prone to inaccuracies and biases. Emerging wearable sensing technology may offer a promising solution. This protocol paper describes a study investigating the physiological responses to energy intake utilising a customised wearable multi-sensor band, which is specifically designed to monitor multiple physiological and behavioural responses related to eating and digestion events for dietary monitoring.

**Methods and Analysis:** This study will recruit 10 healthy volunteers from databases and public advertisements, with informed consent required prior to participation. Participants will attend two main study visits at a clinical research facility, consuming pre-defined high- and low-calorie meals in a randomised order. Wearable sensors will be worn throughout the eating episodes to track hand-to-mouth movements and physiological changes, including skin temperature (Tsk), heart rate (HR), and oxygen saturation (SpO_2_). These sensor readings will be validated against a traditional bedside monitor which additionally measures blood pressure. Blood samples will be collected via intravenous cannula to measure blood glucose, insulin, and hormone levels. Relationship between eating episodes (e.g., occurrence, duration, use of cutlery, high- vs. low-calorie) with hand movement patterns, as well as physiological and blood biochemical responses, will be analysed.

**Ethics and Dissemination:** Ethical approval has been granted by London – Westminster Research Ethics Committee. REC reference: 23/PR/1379. Written informed consent will be obtained from all participants. Findings will be disseminated through peer-reviewed journals, conferences, and seminar presentations. Protocol V.5. Protocol date: 16 January 2024.

**Trial Registration Number:** NCT06398340.

**Strengths and Limitations of This Study:**

- This study is the first trial to develop an objective and reproducible wearable dietary monitoring tool by tracking physiological and motor changes. The novelty of this tool is the ability to estimate food intake without capturing food images which raises fewer privacy concerns compared to the existing technologies in this field.
- The comprehensive monitoring of various physiological and behavioural parameters provides a thorough evaluation of the relationship between eating behaviour and physiological changes, with validation through bedside monitors.
- The exploratory outcomes of the study offer the opportunity to identify physiological markers that can be tracked by a wearable device to predict postprandial blood glucose and hormone levels that have implications for glycaemic control and appetite regulation.
- This study recruits healthy subjects, and investigations should be expanded to include disease groups such as obese individuals who may have different cardiac outputs.
- The study will be conducted under a controlled environment and requires further investigations in real-life settings.

## Introduction

Assessing dietary intake across populations is a fundamental challenge in nutrition research. Traditional dietary assessment methods, such as food diaries, rely on self-reporting of individuals and suffer from poor accuracy and recall bias. Food records are estimated to cause 11-41% underestimations for energy intake ^1^. Furthermore, traditional methods are labour- and time-intensive and require participants to manually record their food intake ^2^. Adherence to dietary self-reporting is challenging even with mobile-based applications e.g., MyFitnessPal ^3^.

Wearable sensing technology is emerging to ease the procedure of dietary assessment. An effective technique is the wearable system based on inertial measurement units (IMUs), which capture eating gestures via monitoring wrist motions that correspond to the hand-to-mouth movements during eating ^4,5^. The advantages include its reliability and capability to capture eating episodes and detect the time, speed, and duration of eating. However, the system relying solely on IMUs is unable to provide information on energy intake. Camera-based sensors could provide measurements of food volume consumption ^6^; however, this method requires complex image processing and raises some privacy concerns. To date, there is a lack of wearable tools that measure physiological responses to food consumption. Such information is essential for determining the interaction between diet and health.

Food intake and digestion increase metabolism, body temperature and intestinal oxygen consumption. Elevated skin temperature (Tsk), increased heart rate (HR) and lowered blood oxygen saturation (SpO_2_) were observed during or after meals ^7–10^; the increase of HR was found to be significantly correlated to meal size (r = 0.990; P = 0.008) in six healthy volunteers ^11^. This raises the possibility of using these physiological responses as potential indicators of food and energy intake ^12^.

A significant challenge in developing applications that use physiological responses to monitor food intake is that a single physiological parameter like HR, can be influenced not only by food and digestion, but also by confounding factors, including exercise. To address this, this study aims to integrate physiological parameters with existing motion sensors—such systems that have already demonstrated high accuracy in distinguishing eating events from other activities in real-life environments ^13^. We hypothesize that combining multiple physiological parameters with existing motion sensors could provide an objective tool for detecting eating events and estimating food consumption.

## Aims and Objectives

This study aims to investigate the relationships between physiological responses and food intake via a customised wearable monitor with non-intrusive sensors. This system integrates commercially available, cost-effective sensors and products to measure diet-related physiological parameters. Our findings could accelerate the development of objective wearable tools for dietary assessment, benefiting both nutrition research and dietary self-monitoring in the future.

**Primary objective**: to investigate the changes in heart rate associated with dietary events (pre- vs post-meal) and energy loads (high vs. low-calorie meals).

### Secondary objective

- To investigate the changes of other physiological changes such as skin temperature, oxygen saturation, and blood pressure associated with dietary events (pre- vs post-meal) and energy loads (high vs. low-calorie meals).
- To investigate the changes in eating behaviours by tracking hand movements associated with dietary events (pre- vs post-meal) and energy loads (high vs. low-calorie meals).

### Exploratory objective

- To explore the relationship between physiological features (heart rate, skin temperature, oxygen saturation and blood pressure) with glycaemic biomarkers, including blood glucose levels, insulin levels, and hormonal levels.

## Methods and Analysis

### Study Setting and Participants

The study will be conducted at the National Institute for Health and Care Research (NIHR) Imperial Clinical Research Facility. Participants will be monitored throughout the interventions, ensuring adherence to the study protocol.

#### Sample size

We plan to recruit ten healthy male and female participants in total. As described previously, the primary outcome is the heart rate change before and after meals. No similar study measured heart rate using a wearable system following meals; however, Sidery & Macdonald ^9^ investigated the heart rate responses in eight healthy subjects via a traditional device and reported significant differences between high and low-energy meals. A power analysis based on this study ^9^, with an effect size (d=1.29), an alpha of 0.05, and a targeted power of 0.9, the analysis suggests that a minimum of 9 participants would be adequate to detect significant heart rate differences.

For behavioural analysis with IMU, previous studies have demonstrated that high accuracies have been achieved with similar sample sizes to our study. For example, a benchmarked dataset with recordings of 18 activities performed by 9 subjects was provided for human activity monitoring. According to the results, several classification models achieved accuracies higher than 95% ^14^. Another study with 10 subjects shows an accuracy of 97.07% in IMU-based activity recognition^15^.

#### Inclusion Criteria

- Male or female
- Age between 18-65 years (inclusive)
- Body mass index (BMI) of 18-30 kg/m^2^
- Willingness and ability to give written informed consent
- Willingness and ability to understand, to participate and to comply with the study requirements

#### Exclusion Criteria

- Outside of the specified age and BMI range
- Chronic medical conditions including eating disorders, diabetes, obesity, hypertension, cancer, acute infectious disease, renal disease, cardiovascular disease, and chronic gastrointestinal condition
- Taking part in another research study or donating any blood in the last 3 months

#### Withdrawal Criteria

Each participant has the right to withdraw from the trial at any time without giving a reason. In addition, the investigator may discontinue a participant from the trial at any time if the Investigator considers it necessary for any reason including:

- An adverse event which requires discontinuation of the trial application or results in inability to continue to comply with trial procedures.
- Loss of capacity to consent.

### Menu Design

Participants will consume a high- and a low-calorie meal as shown in Table 1. These meals have been chosen to represent commonly consumed food choices in the Western diet. Foods will be purchased from local supermarkets in London. The high-calorie meal contains 1052 Kcal while the low-calorie contains 301 Kcal. Our unpublished meta-analysis suggests that such energy disparity would result in significantly different HR increases, regardless of food form or nutritional compositions. Eating instructions will be provided on the use of cutlery and hands to reflect the common eating behaviours in real-world settings.

**Table 1.** Nutrient Composition of High- and Low-Calorie Meals. This table presents the percentages of carbohydrates (CHO), protein, and fats in meals categorised as high and low in calories. Each row details the macronutrient distribution for individual meals, accompanied by specific eating instructions.

| **Eating Instruction** | **Meal Content** | **Weight/ g** | **Energy /kCal** | **CHO/ g (%)** | **Protein /g (%)** | **Fat/ g (%)** |
| --- | --- | --- | --- | --- | --- | --- |
|  | **High Calorie** | **365** | **1052** | **124.8 (48.9)** | **39.7 (15.6)** | **42 (37.0)** |
| No cutlery, use both hands | Sainsbury's Margherita Pizza | 265 | 724 | 90.6 (50.1) | 35.0 (19.3) | 23.0 (28.6) |
| With cutlery, use one hand | Sainsbury's New York Cheesecake | 100 | 328 | 34.2 (41.7) | 4.7 (5.7) | 19.0 (52.1) |
|  | **Low Calorie** | **380** | **301** | **28.8 (38.3)** | **19.2 (25.5)** | **11.65 (34.8)** |
| With cutlery, use both hand | Sainsbury's Chicken Caesar Salad | 180 | 215 | 10.6<br>(19.7) | 17.8<br>(33.1) | 10.8<br>(45.2) |
| No cutlery, use one hands | Sainsbury's Fruit Bowl | 200 | 86 | 18.2<br>(84.7) | 1.4<br>(6.5) | 0.85<br>(8.9) |

### Wearable Sensors and Data Acquisition

A customised multi-sensor wristband is used as a wearable monitor of physiological responses and behavioural data (Figure 1). Participants will wear the multi-sensor monitor on the wrist 5 minutes before meal consumption and up to 1 hour post-prandial. This monitor will be equipped with multiple sensors designed to record various physiological responses and physical activities potentially relevant to food intake. The functional sensors in the wearable wristband include:

- **An integrated pulse oximeter module** for automatic tracking of HR and blood SpO2 levels. With signal process algorithms embedded, the module will provide digital readings of HR and SpO_2_ levels.
- **A photoplethysmography (PPG) sensor module** for recording continuous traces of blood volume changes, can reflect HR and additional cardiorespiratory information.
- **A skin surface temperature sensor** for continuously monitoring Tsk variation when the band is wearing.
- **A movement measurement sensor module**, specifically, an Inertial Measurement Unit (IMU), including an accelerometer, a gyroscope and a magnetometer, to record and analyse eating behaviours.
- **A flexible force sensor** for monitoring variations in tightness when worn. This ensures proper tension on the wristband and secure contact of other sensors (such as PPG sensor) to the participants’ skin.

**Figure 1.**
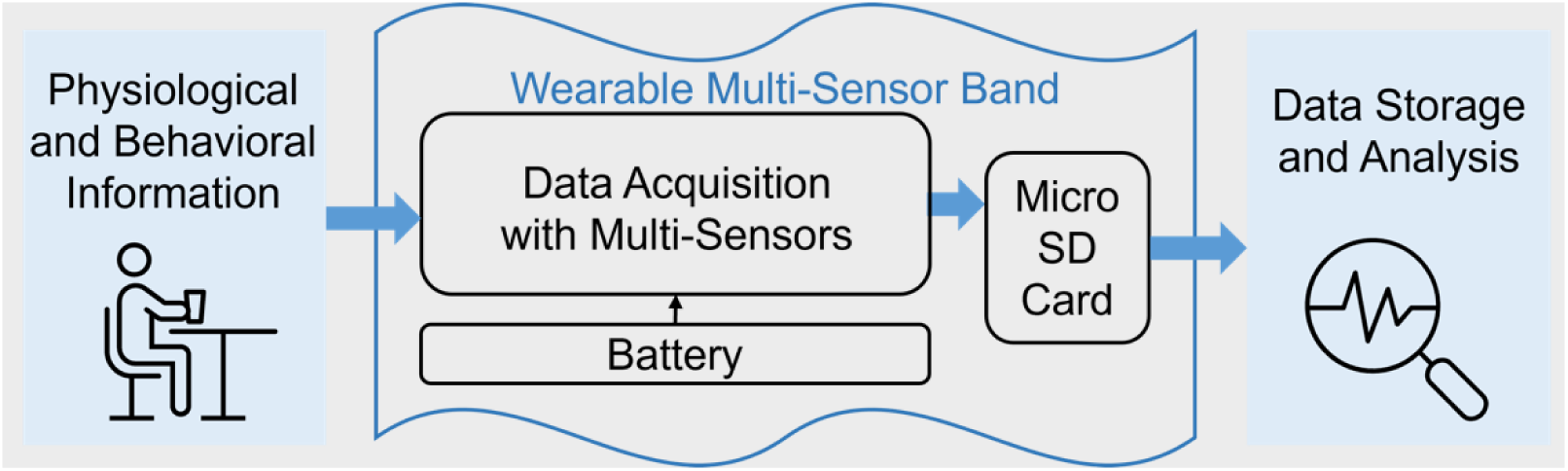
Wearable multi-sensor band for physiological and behavioural monitoring.

Table 2 shows various sensors and their respective measurements, along with how these measurements correlate to the status of food intake or wearable monitor. In conjunction with the wearable multi-sensor monitor, a bedside vital sign monitoring system will also be utilised. This system will measure blood pressure and provide additional validation data on blood SpO_2_ levels and HR via a pulse oximeter.

**Table 2.**
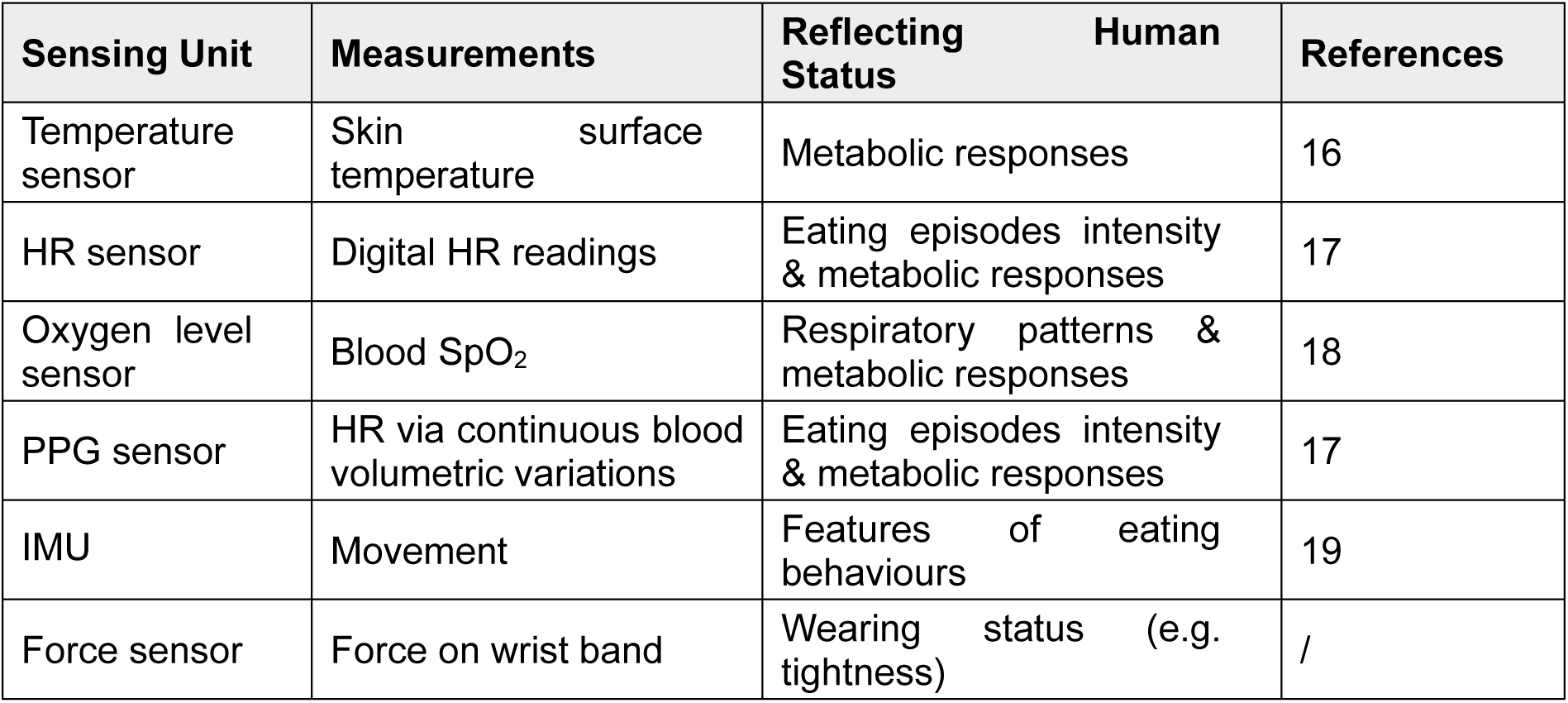
Overview of Sensing Equipment and Corresponding Measurements for Monitoring Dietary Intake.

### Study Procedures

A flowchart outlining study procedures, including recruitment, pre-screening, obtaining consent, study visits, data collection and analysis is shown in Figure 2.

**Figure 2.**
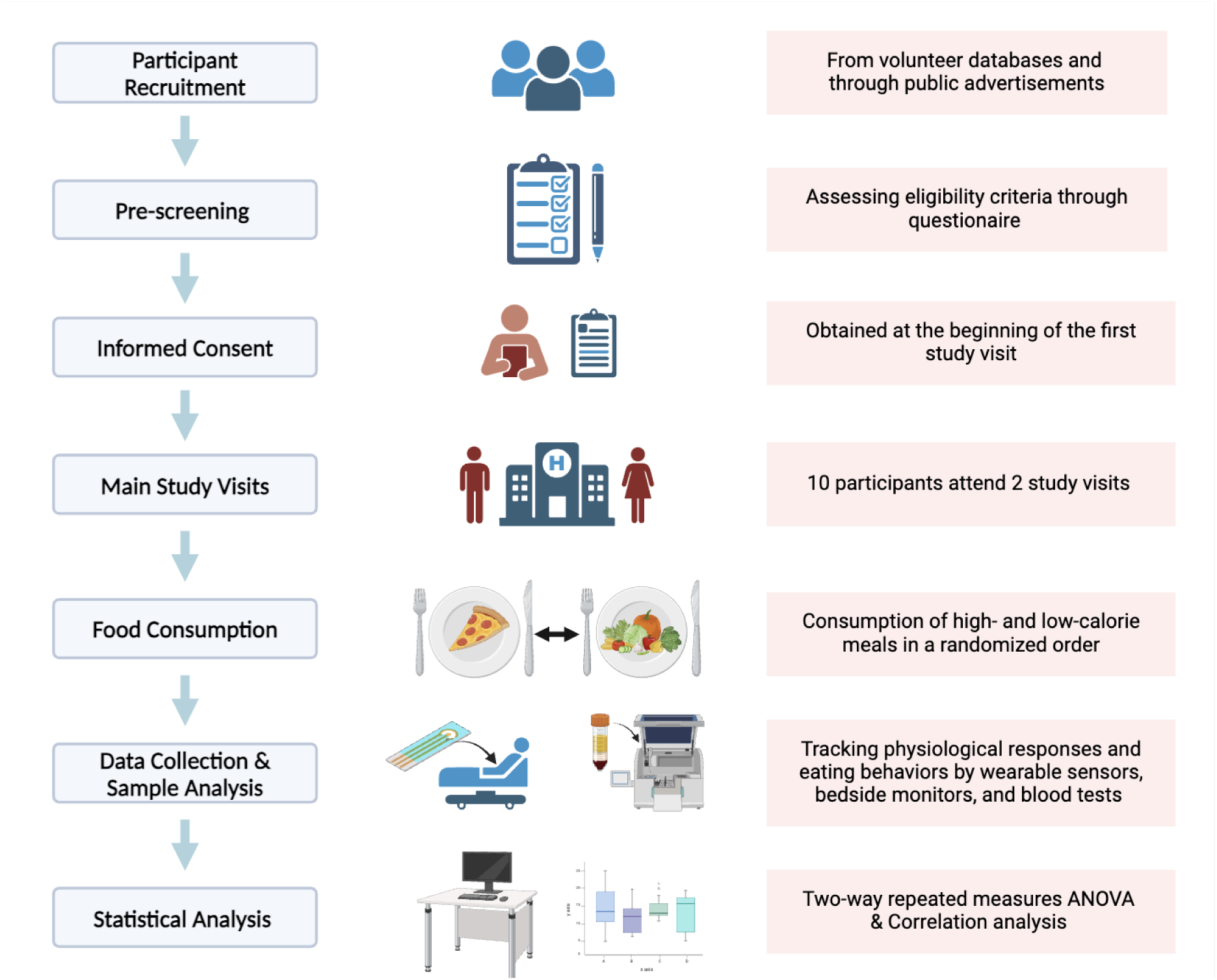
Flowchart of clinical study design. This flowchart outlines the process of investigating the effects of high- and low-calorie meal consumption on physiological responses. The study design involves the following steps: Participant Recruitment, Pre-screening, Informed Consent, Main Study Visits, Food Consumption, Data Collection, Sample and Statistical Analysis.

#### Participant Recruitment

Participants will be recruited from healthy volunteer databases and potential participants will also be able to contact the research team through information provided in advertisements placed in newspapers and public buildings.

#### Pre-Screening

Inclusion and exclusion criteria will be assessed via a questionnaire. Data from ineligible participants will not be retained.

#### Informed Consent

Study procedures will be explained by study team who has received training. Consents will be obtained at the beginning of the first study visit.

#### Randomisation

The sequence of receiving high- and low-calorie meals will be randomised by an independent researcher using the ‘sealed envelope’ website.

#### Blindness

Blinding is not possible for researchers and participants due to the distinct nature of high-calorie and low-calorie diets.

#### Pre-visit

The day before the study visit, participants will be requested to refrain from strenuous exercise and alcohol and to arrive having fasted from the evening before.

#### Study Visit Overview

The overview of the study visit is shown in Figure 3. Participants will attend two main study visits at the NIHR Imperial Clinical Research Facility, during which they will consume high and low-calorie meals in a randomized order. Each visit will involve monitoring physiological parameters such as Tsk, blood SpO2, and HR using a bedside monitor and a non-invasive, wearable multimodal sensor band developed by Dr Mayue Shi. Each study visit will take no more than 2 hours.

**Figure 3.**
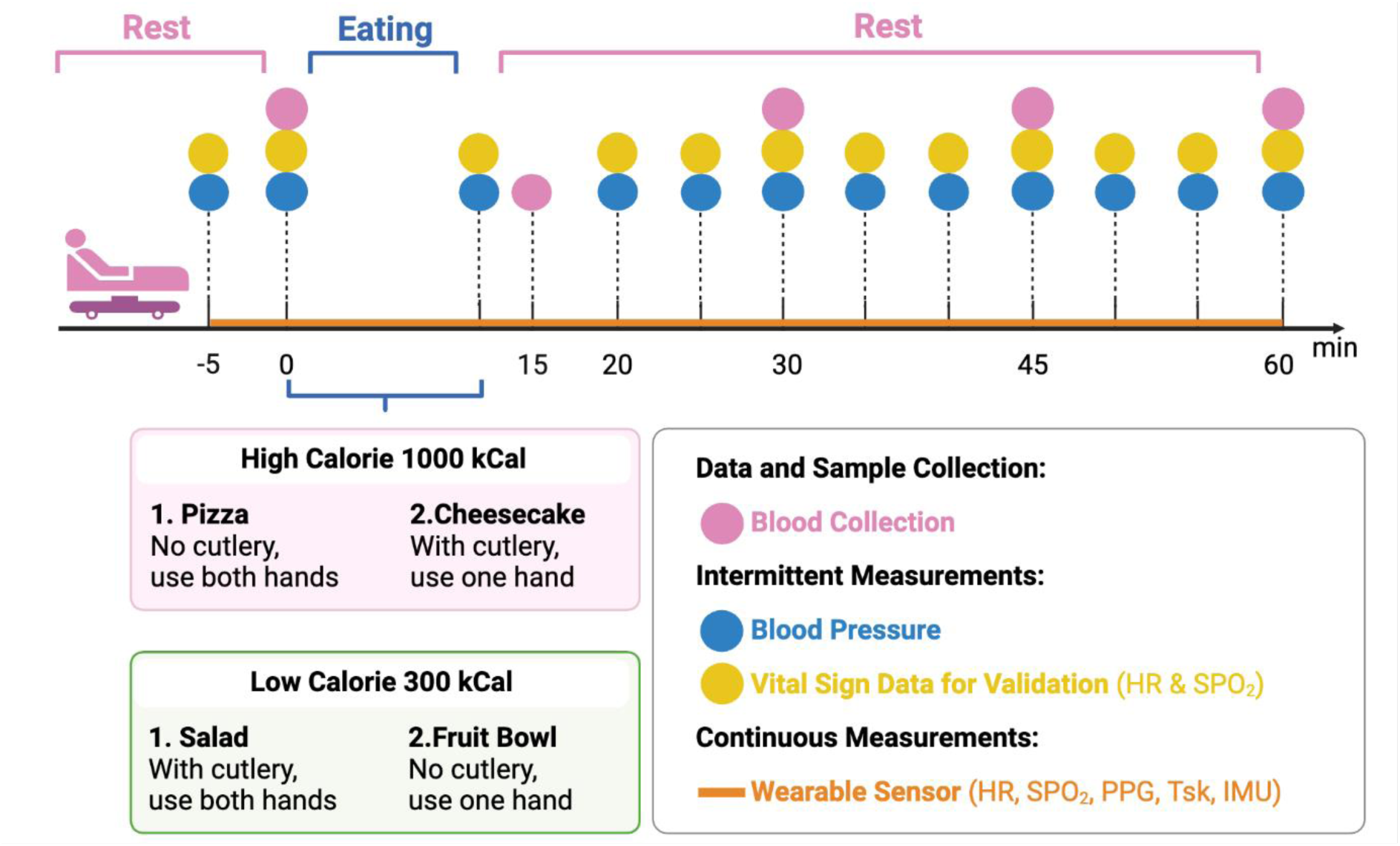
Overview of Study Timeline and Data Collection. Participants will undergo a 5-minute pre-prandial period and a 60-minute eating and postprandial period. High-calorie meals include pizza (eaten without cutlery) and cheesecake (eaten with cutlery), while low-calorie meals include salad (eaten with cutlery) and a fruit bowl (eaten without cutlery). Wearable sensors (orange line) will take continuous measurements of HR, SpO2, PPG, Tsk, and IMU data. Blood pressure and vital signs (blue and yellow circles) will be measured intermittently by a bedside monitor. Blood collection (pink circles) will be collected at designated intervals. Abbreviation: HR: Heart Rate; IMU: Inertial Measurement Unit; PPG-Photoplethysmography; SpO_2_: Oxygen Saturation; Tsk: Skin Temperature.

#### Body Measurement

Participant characteristics including weight, height, BMI, waist circumference, and body composition will be measured.

#### Preparation

Participants will receive thorough explanations of the study procedures and necessary precautions at each visit. Precautions include emptying the bladder, proper seating with the arm at heart level, removing clothing on the left arm for the blood pressure cuff, ensuring proper cuff fit, refraining from leg crossing, and maintaining silence during measurements. A preparation checklist (Appendix 1) will ensure all steps are completed before starting the study. The study’s detailed tick box timeline will also ensure that all actions, from device setup to post-study procedures, are executed accurately and timely. The wearable band will be pre-tested by connecting it to a laptop, to verify that the status of each sensor is correct before the study begins.

#### Device Setup and Measurements

Five minutes before the study begins, a wearable sensor will be placed on the participant’s right arm, which remains in place for about 60 minutes for continuous monitoring. Concurrently, a pulse oximeter and a blood pressure cuff, connected to a bedside monitor will be worn for two baseline measurements before the meal. After meal consumption, the pulse oximeter and blood pressure cuff will be continuously worn till the end. Readings at pre-specified intervals (see Figure 3) will be recorded. Please refer to the Appendix x for the data collection form.

#### Dietary Intervention

Participants will be instructed to consume food at their normal pace, adhering to guidelines regarding the use of cutlery or hands as outlined in Table 1.

#### Blood Sample Collection

Blood samples will be collected by venepuncture, beginning at baseline before the meal, and continuing every 15 minutes up to 60 minutes post-meal (0, 15, 30, 45, 60 min). At each time point, 5 ml of blood samples will be collected in vacutainers and centrifuged at 2500 g at 4°C for 10 minutes. 500ul of supernatant will be collected into aliquots and stored at -80 °C for further analysis.

#### Post-Study Procedures

After the study, all devices will be removed, and data storage on the wearable sensor will be confirmed.

Figure 3 outlines the protocol for a study designed to compare the physiological effects of consuming high versus low-calorie meals over 60 minutes. The timeline is segmented into rest and eating phases. The high-calorie meals include pizza and cheesecake, while the low-calorie options consist of salad and a fruit bowl, each accompanied by specific instructions for consumption. Continuous physiological monitoring is depicted by a solid orange line, representing the use of a wearable sensor that tracks real-time physiological changes such as HR, SpO_2_ and Tsk throughout the session. Key data collection points on the timeline are marked for clarity: blood samples are collected at intervals indicated by pink circles, blood pressure measurements marked by blue circles, and vital sign data for validating wearable sensor data, specifically SpO_2_ and HR, are monitored and recorded at times shown by yellow circles.

### Outcomes

The primary outcome is change in heart rate associated with dietary events (pre- vs post-meal) and energy loads (high vs. low-calorie meals).

Secondary outcomes include the motor features and physiological responses including skin temperature, oxygen saturation, and blood pressure associated with dietary events (pre- vs post-meal) and energy loads (high vs. low-calorie meals).

The exploratory outcome is the relationship between physiological changes and glycaemic biomarkers, including blood glucose, insulin, and hormonal levels.

### Analysis Plan

#### Wearable Sensor Data Analysis

Data acquired from wearable sensors will be pseudonymised and saved after each visit. Data will be first evaluated with quality assessment. Signals will be assessed using proper techniques, including visualisation, signal-to-noise ratio (SNR) analysis and outlier detection to identify potential noise, environmental interference and missing values. After quality assessment, data from different sensors will be processed respectively. IMU, PPG and Tsk data will be filtered to remove high-frequency noise. Then, data will be analysed with statistical methods to get physiological and behavioural parameters such as HR, and motion intensity during food intake. In addition, machine learning methods would be considered to explore the possibility of tracking food intake behaviours, such as the patterns of eating behaviours.

#### Blood Sample Analysis

Blood samples will be collected for glucose, insulin, and gut hormone measurements. Plasma glucose concentrations will be measured using a GLUC-PAP Kit from Randox (Randox Laboratories Ltd., UK) following the instructions provided. Total GIP and GLP-1 will be measured using human ELISA kits from (Merck, UK).

#### Statistical Analysis

To address the primary outcome, a mixed-effects model will be conducted to analyse the effects of time and meal type (high vs. low-calorie meals) on heart rate.

Similarly, for the secondary outcomes, mixed-effects models will be applied to each physiological parameters SpO_2_, Tsk, and blood pressure.

For exploratory outcomes, correlation analysis will be conducted to investigate the possible associations/trends between physiological changes and blood glycaemic responses. The exploratory outcomes are intended to understand possible trends and associations, which may require further validation with a larger cohort.

Missing data: Mixed-effects models use all available data points and do not require missing data imputation.

This protocol was developed in accordance with the SPIRIT reporting guidelines^20^.

## Data Management and Safety

### Data Handling and Storage

Data will be managed in strict compliance with data protection regulations and ethical guidelines to ensure integrity and confidentiality.

#### Confidentiality Measures

***1. Pseudonymisation:*** Participants’ personal information will be pseudonymised. Each participant will be assigned a unique ID linked to their consent forms, which include names and study codes. These forms will be securely stored in locked cabinets within Imperial College London’s Investigative Medicine Section.
***2. Secure Storage:*** Participants’ contact details will be stored on a password-protected drive, accessible exclusively to the research team. This measure ensures that personal data remains confidential and is only available to authorized personnel.
***3. Electronic Data Security:*** All electronic data will be protected using strong password protocols. Study data will be referenced only by study codes, except for specific instances accessible to authorized researchers, thereby maintaining pseudonymisation across all digital records.

#### Data Monitoring

The study will be subject to audit by Imperial College London under their remit as sponsor and other regulatory bodies to ensure adherence to GCP and the UK Policy Framework for Health and Social Care Research.

## Patient and Public Involvement

Four public contributors were invited through Imperial College Healthcare NHS Trust to review participant information sheets. Feedback was collected to improve wording and help translate scientific terms into plain language that enhances communication with participants.

## Ethics and Dissemination

The study will be conducted in accordance with the recommendations for physicians involved in research on human subjects established by the Declaration of Helsinki ^21^. The protocol has been approved by the London – Westminster Research Ethics Committee and the Health Research Authority (HRA) (reference no: 23/PR/1379). Additionally, each participating NHS Trust will confirm its capacity and capability before the study can enrol participants or initiate any research activities. Review committee will be noticed and give approval for substantive protocol amendments.

Each participant must provide consent to join the study only after receiving a comprehensive explanation, an information leaflet, and adequate time to consider their decision. Consent will be documented with a signed form. Participants have the right to decline participation or withdraw from the study without providing reasons.

The findings of the research will be presented at scientific conferences and published in an open-access, peer-reviewed journal. In addition, we will be collaborating with patient groups and professional groups to disseminate the findings via multiple media channels such as patient association publications, print and broadcast media. No participants’ identifiable data will be included.

## Discussion

Unhealthy diets are a major contributor to the global non-communicable disease (NCD) burden^22^. However, the lack of accurate tools to collect dietary information limits our understanding between diet and health and disables healthcare professionals from providing the most effective dietary advice.

This study will improve understanding of how a wearable device incorporating physiological and behavioural monitoring can track eating behaviours and food intake. This will address current limitations of wearable sensors that most types can detect eating events but fail to estimate food portions ^4^. The innovation also lies in the development of sensors that are less costly and raise fewer ethic concerns compared to the existing technologies in this field such as camera-based systems ^23^. This offers a great potential for further development of smartwatch products which are widely accepted by the public for health monitoring.

The expected outcomes will have an impact across various domains. Academically, the proposed wearable sensors may provide an objective tool for estimating energy intake, allowing deeper and more accurate investigations into the interactions between diet and health. Our outputs will have social and economic impacts on stakeholders in the food system, healthcare, and patient groups. The sensors and algorithms in development might enable individuals to monitor their physiological and food intake status in real time. This will be most beneficial for obese patients to improve their self-management of energy intake. The technology may be implemented in collaboration with healthcare professionals, improving communication with patients about their habitual dietary intake and enabling more accurate treatment. The sensors may be applied to biopharmaceutical and healthcare sectors to simplify dietary data collection procedures. Enhanced monitoring of the health status of individuals can reduce the economic burden on the NHS.

The findings shall be interpreted in consideration of these limitations of the study design. First, the study will test two common and contracting meals representing a healthy versus an unhealthy dietary pattern; however, the meal design has not incorporated the specific dietary requirements such as vegetarian or vegan options. A wilder range of test meals should be investigated to generalise current results and test them in real-life settings. In addition, the current commercial sensor modules have not been tested specifically for food intake monitoring before. The most convincing and optimised sampling frequency and magnitude ranges of each sensor for food intake monitoring remain unclear. Considering the data acquisition board’s performance and data quality, the maximum sample frequency was set to 50 Hz. It is needed to evaluate the sensor modules’ performance in food intake monitoring.

In conclusion, this paper presents a protocol aimed at developing a wearable, automatic dietary monitoring tool based on multiple physiological and behavioural signals. Such a tool, if proven feasible, could lead to significant progress in nutritional management of NCDs by providing objective dietary assessments and therefore, more efficient interventions to control these diseases.

## List of Abbreviations

BMI: Body Mass Index
CHO: Carbohydrates
HR: Heart Rate
IMU: Inertial Measurement Unit
NCD: Non-Communicable Disease
PPG: Photoplethysmography
SNR: Signal-to-Noise Ratio
SpO_2_: Oxygen Saturation
Tsk: Skin Temperature

## Declarations

### Patient Consent for Publication

Not required.

### Authors’ Contributions

M.C. and M.S. designed and directed the project; M.C. and M.S. acquired funding as Co-PIs; M.S., J.Z., and M.C. wrote this protocol.

## Supporting information

Participant Consent Form

SPIRIT Checklist

## Data Availability

All data produced in the present study will be available upon reasonable request to the authors

## Appendix

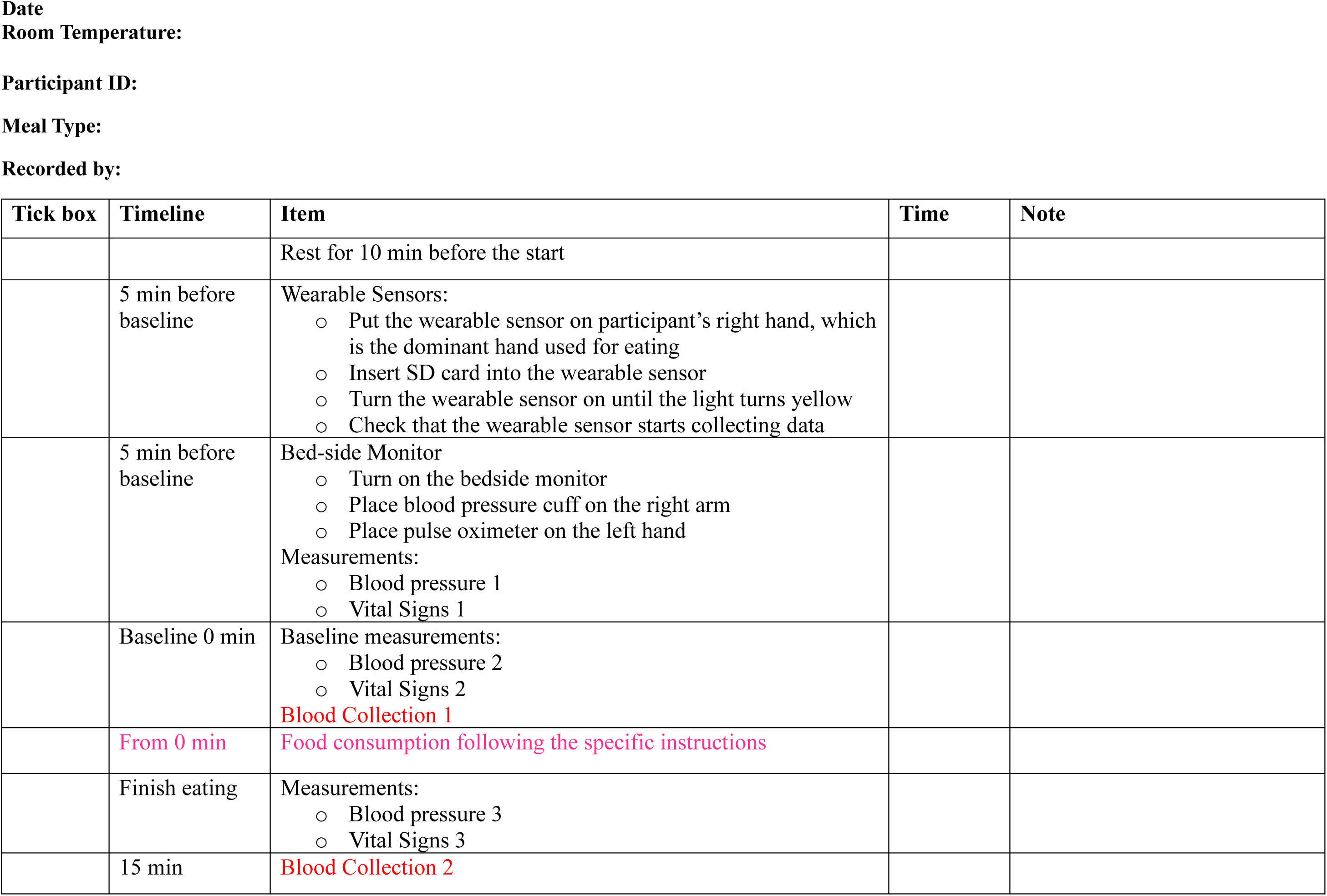
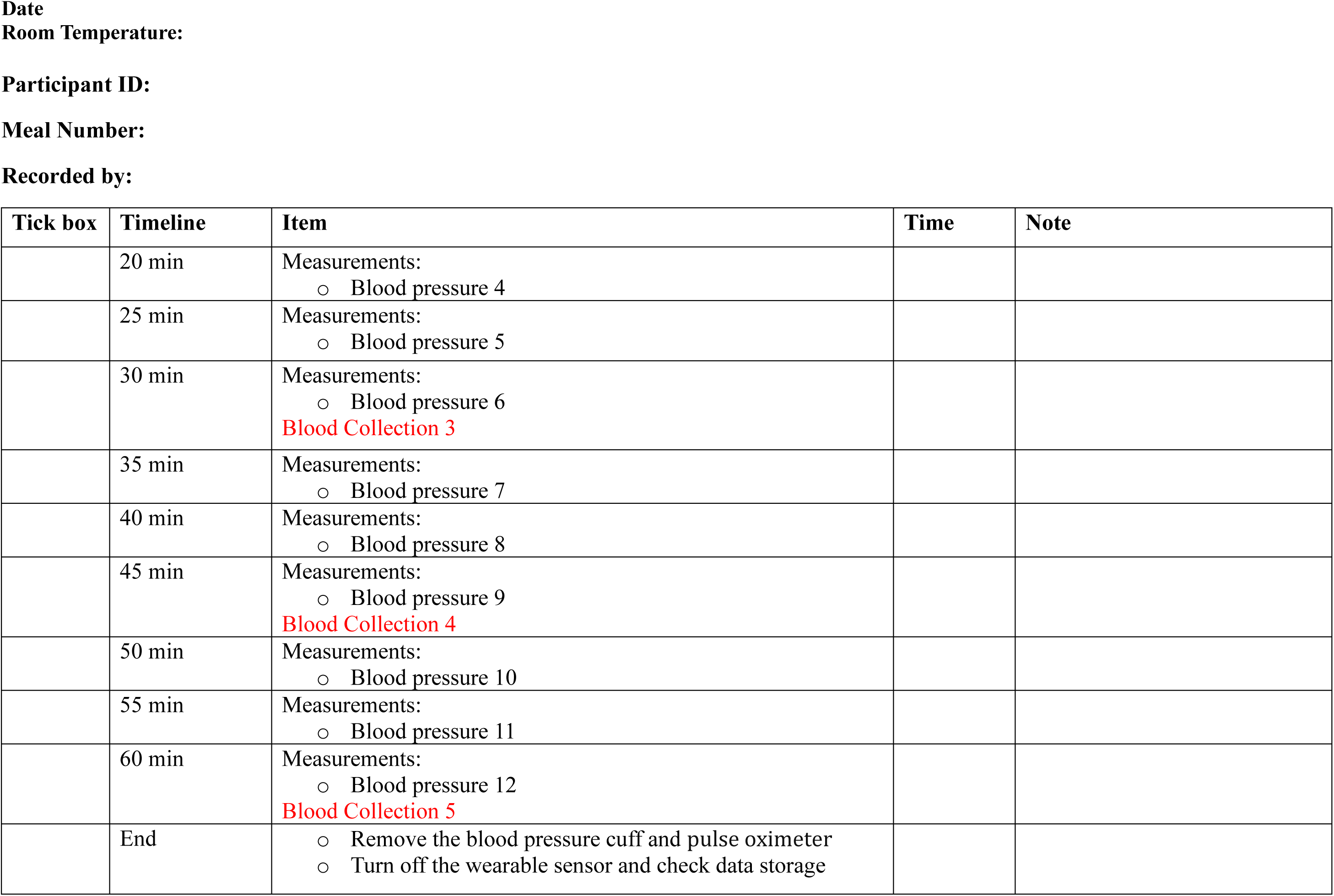
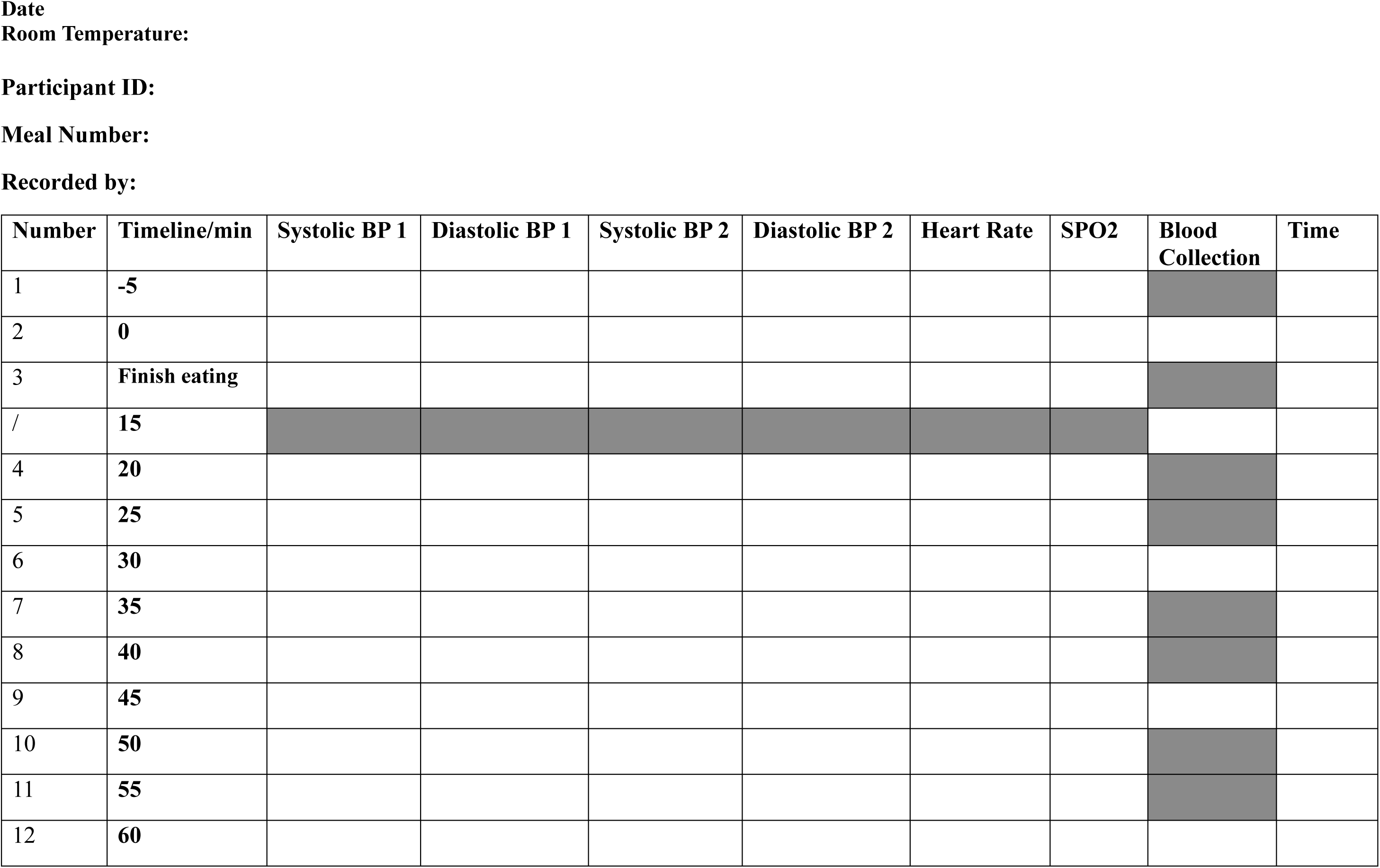

