## Supplementary material for "Multiple Physiological and Behavioural Parameters Identification for Dietary Monitoring Using Wearable Sensors: A Study Protocol": Participant Consent Form

Imperial College London

### Identifying Physiological Markers for Monitoring Dietary Behaviours

Participant Consent Form: Version4_Date 16/01/2024

Name of Principal Investigator: Dr Mingzhu Cai

***Please initial the box***

| |
|---|
| 1. I confirm that I have read and understand the participant information sheet version dated for the study entitled *"Identifying Physiological Markers for Monitoring Dietary Behaviours"* and have had the opportunity to ask questions which have been answered fully. |
| 2. I understand that my participation is voluntary, and I am free to withdraw at any time, without giving any reason and without my legal rights nor treatment / healthcare being affected. |
| 3. I understand that sections of any of my medical notes may be looked at by responsible individuals from Imperial College London, from NHS Trust or from regulatory authorities where it is relevant to my taking part in this research. |
| 4. I understand that samples and data collected from me are a gift donated to Imperial College. I will not personally benefit financially if this research leads to an invention and/or the successful development of a new test, medication treatment, product or service. |
| 5. I consent to take part in the study entitled *"Developing Physiological Markers and Tools for Monitoring Dietary Behaviours."* |
| 6. I [give/do not give] consent for **my information** to be used to support other research or in the development of a new test, medication, medical device or treatment (mark as applicable) by an academic institution or commercial company in the future, including those outside of the United Kingdom. |
| 7. I [give / do not give] consent for **my samples** collected to be used to support other research or in the development of a new test, medication, medical device or treatment (mark as applicable) by an academic institution or commercial company in the future, including those outside of the United Kingdom. |
| 8. I [give / do not give] consent to being contacted about the possibility to take part in other research studies. |
| 9. I [agree / I do not agree] to my tissue samples being used to undertake genetic research which may have the potential to generate data that can be tracked back to me. |
| 10. I [agree / I do not agree] that the research team informs my GP of my participation in the study. |
| 11. I would like to receive study results via [email / phone] at the end of the study. |
| 12. I [ would / would not] like to receive a copy of the study results once published. |

_________________________ _________________ _________________

Name of Subject Signature Date

_________________________ _________________ _________________

Name of Person taking consent Signature Date

1 copy for participant; 1 copy for Principal Investigator,1 copy for hospital notes

To ensure confidence in the process and minimise risk of loss, all consent forms <u>must</u> be printed, presented and stored in double sided format
